# Profiling the positive detection rate of SARS-CoV-2 using polymerase chain reaction in different types of clinical specimens: a systematic review and meta-analysis

**DOI:** 10.1101/2020.06.11.20128389

**Authors:** George M. Bwire, Mtebe V. Majigo, Belinda J. Njiro, Akili Mawazo

## Abstract

**Background:** Testing is one of the commendable preventive measures against coronavirus disease (COVID-19), and needs to be done using both most appropriate specimen and an accurate diagnostic test like real time reverse transcription polymerase chain reaction (qRT-PCR). However, the detection rate of severe acute respiratory syndrome coronavirus 2 (SARS-CoV-2) RNA from different clinical specimens after onset of symptoms is not yet well established. For guiding the selection of specimens for clinical diagnosis of COVID-19, a systematic review aiming at profiling the positive detection rate from different clinical specimens using PCR was conducted.

**Methods:** The systematic search was done using PubMed/MEDLINE, Science direct, Google Scholar, among others. The search included studies on laboratory diagnosis of SARS-CoV-2 from different clinical specimens using PCR. Data extraction was done using Microsoft Excel spread sheet 2010 and reported according to PRISMA-P guidelines. Using Open Meta Analyst software, DerSimonian–Laird random effects analysis was performed to determine a summary estimate (positive rate [PR]/proportions) and their 95% confidence interval (95%CI).

**Results:** A total of 8136 different clinical specimens were analyzed to detect SARS-CoV-2, with majority being nasopharyngeal swabs (69.6%). Lower respiratory tract (LRT) specimens had a PR of 71.3% (95%CI:60.3%-82.3%) while no virus was detected from the urinogenital specimens. Bronchoalveolar lavage fluid (BLF) specimen had the PR of 91.8% (95%CI:79.9-103.7%), followed by rectal swabs, 87.8 % (95%CI:78.6%-96.9%) then sputum, 68.1% (95%CI:56.9%-79.4%). Low PR was observed in oropharyngeal swabs, 7.6% (95%CI:5.7%-9.6%) and blood samples, 1.0% (95%CI: -0.1%-2.1%), whilst no SARS-CoV-2 was detected in urine samples. Nasopharyngeal swab, a widely used specimen had a PR of 45.5% (95%CI:31.2%-59.7%).

**Conclusion:** In this study, SARS-CoV-2 was highly detected in lower respiratory tract specimens while there was no detected virus in urinogenital specimens. Regarding the type of clinical specimens, bronchoalveolar lavage fluid had the highest positive rate followed by rectal swab then sputum. Nasopharyngeal swab which is widely used had a moderate detection rate. Low positive rate was recorded in oropharyngeal swab and blood sample while no virus was found in urine samples. More importantly, the virus was detected in feces, suggesting SARS-CoV-2 transmission by the fecal route.

## Introduction

Coronavirus diseases 2019 (COVID-19) is a highly infectious and an emerging respiratory disease caused by a severe acute respiratory syndrome coronavirus 2 (SARS-CoV-2) (1). Although its pathogenesis is still unclear but the current evidence associated SAR-CoV-2 infection with angiotensin converting 2 receptors (2–4). On the other hand, real-time reverse transcription– polymerase chain reaction (qRT-PCR) of upper respiratory specimens, mainly nasopharyngeal swabs has been widely used to confirm the clinical diagnosis of COVID-19 (5).

However, there are reports of SARS-CoV-2 detections from other sites including feces (6,7), and therefore extending the spectrum of specimens other than those from respiratory tract to be considered for clinical diagnosis of SARS-CoV-2. From the beginning, health authorities, i.e., World Health Organization (WHO) (8) and Center for Disease Prevention and Control (CDC) advocated on massive and rapid testing of COVID-19. Testing as one of commendable approaches in the fight against COVID-19 pandemic needs be done using both most appropriate specimen (6) and an accurate diagnostic test like PCR (9). If testing is properly done especially during this time when the re-opening is on its way in most of the countries, the risk of SARS-CoV-2 transmission will be minimized.

Currently, the detection profile of SARS-CoV-2 RNA from different clinical specimens using qRT-PCR after onset of symptoms is not yet well established. Recent study by Wang et al (6) using 1070 clinical specimens such as bronchoalveolar lavage fluid (BLF), fibrobronchoscope brush biopsy (FBB), sputum, nasal and pharyngeal swabs, urine, feces and blood collected from 205 patients revealed a dynamic profile with high detection rate of virus from lower respiratory tract (LRT) specimens, i.e., BLF and zero detection from urogenital tract specimen, i.e., urine. Therefore, the aim of this systematic review was to establish the profile of detecting SARS-CoV-2 from different types clinical specimens using a standard diagnostic test (qRT-PCR).

## Methods

### Protocol development

A systematic review protocol was developed based on the question “What is the positivity rate for SARS-CoV-2 using qRT-PCR in different types of clinical specimens”. The review was developed in accordance to Preferred Reporting Items for Systematic Reviews and Meta-Analyses Protocols (PRISMA-P) guidelines (10). The protocol was registered in International Prospective Register of Systematic Reviews (PROSPERO database: https://www.crd.york.ac.uk/PROSPERO with a registration number CRD42020189107).

### Search strategy

A rigorous systematic search strategy was developed with the help from librarian using published guidelines of the Cochrane Collaboration (11). A systematically search from PubMed/MEDLINE, Science Direct and Google scholar (12) was conducted. We also searched the websites of key healthcare organizations such as World Health Organization, Centre for Disease Control and Prevention (CDC). With a help of Google, a supplementary search was done from grey literature sources, pre-prints, journal’s website (JAMA, Lancet, Nature Research and NEMJ). Data from December 31, 2019 onward conducted in human beings and published in English language qualified for inclusion. The strategy was primarily developed for PubMed using keywords (Additional file 1). The search terms were combined using Boolean logic ‘OR’ for synonymous terms and ‘AND’ across elements of PCO (population, comparability and outcome). Filters were set to exclude non-human studies, limit the publication period exclude review and case report articles, among others. Keywords such as “laboratory diagnosis”, “polymerase chain reaction”, “clinical sample”, “clinical specimen”, “novel coronavirus 2019”, “SARS-CoV-2”, COVID-19 were used. This search strategy was adapted to the other databases search. All searched articles from different database were exported into End Note software version X7 (Thomson Reuters, 2015) where duplicates were identified and removed. The articles were grouped into relevant categories as indicated in the PRISMA flow diagram (Fig. 1).

**Figure 1:**
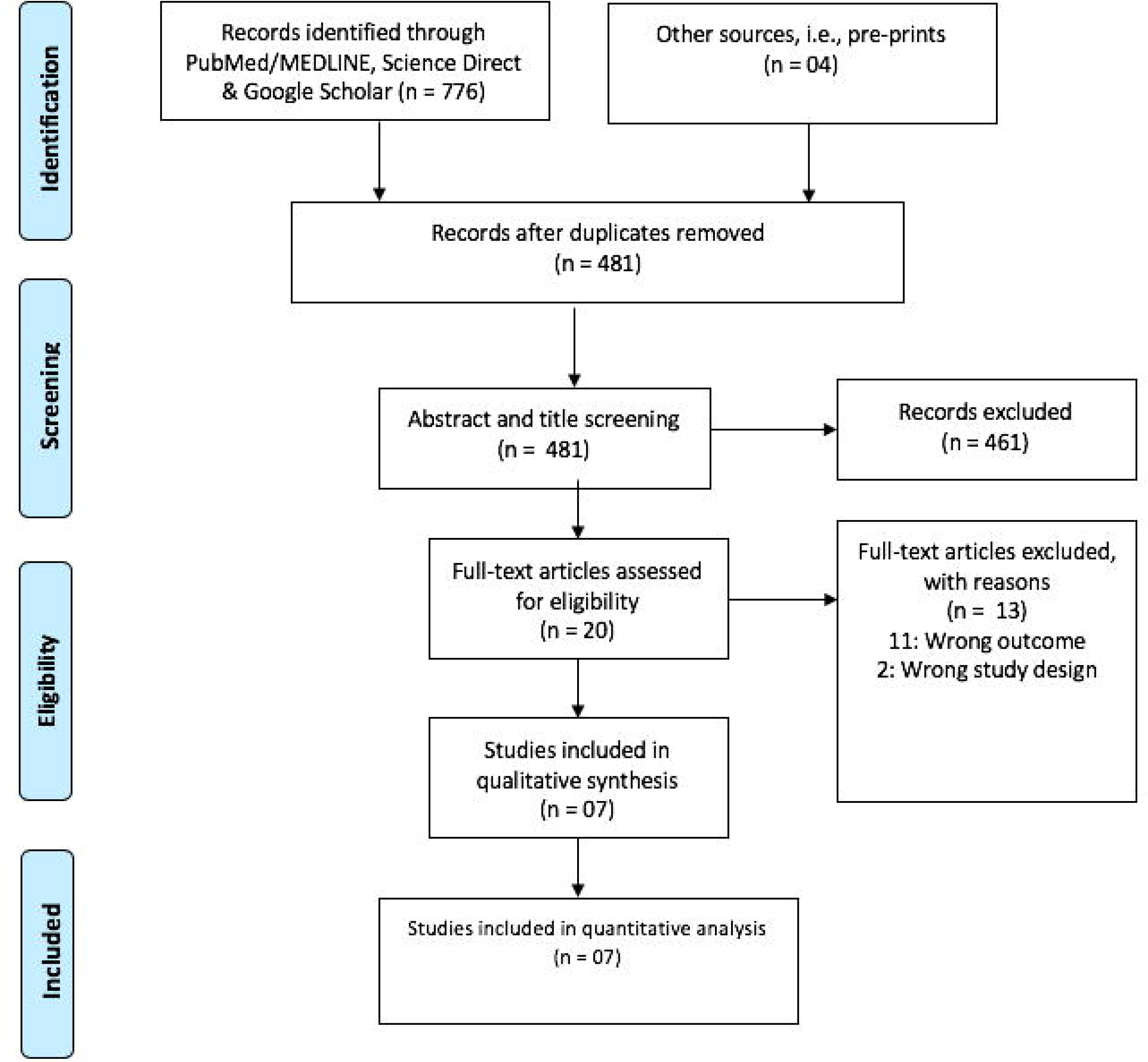
Prims flow chart showing study screening

### Eligibility criteria

Both clinical trials and observational studies (including cross sectional, retrospective and prospective studies) were eligible. Additionally, case series were included, however, considering at least one patient per study, case reports were excluded. This review focused on studies which reported RNA extracted from clinical specimens and determined by qRT-PCR targeting the open reading frame lab gene of SARSCoV-2. A cycle threshold value less than 40 was regarded as positive for SARS-CoV-2 RNA. In this case, the less the cycle threshold value the higher the viral load. Studies which were conducted to determine the diagnostic accuracy, reviews and non-human articles were excluded. For inclusion in the final analysis, at-least 2 samples were required to be reported per study.

### Data extraction

Study selection was managed using EndNote software version X7 (Thomson Reuters, 2015) where two independent reviewers (GMB & BJN) evaluated articles for potential inclusion by screening titles and abstracts followed by full-text screening to determine eligibility for final inclusion. Discrepancies were resolved by consensus, and/or consulting a third reviewer where necessary. Data extracted from study documents, included information about author, year of publication, study design and positivity rate (positive/ total specimen tested) (Table 1). Unavailable, unclear information and additional details were requested from the corresponding author. In some of the studies (7,18,19), patient was regarded as positive, if one of the specimens tested positive. Additionally, at-least 2 specimen reported per test qualified for analysis where a positive patient was considered when one of the specimens tested positive, and recovered patient was considered when at least 2 qRT-PCR consecutive tests tested negative in all tested specimen. The number of laboratory tests were counted based on the specimen tested not number of patients.

**Table 1:** Characteristics of the reviewed studies

| **Author** | **Design** | **Type of specimen** | **Positive test** | **Total test** |
| --- | --- | --- | --- | --- |
| Wang et al.,<br>2020 (6) | Cross sectional | Bronchoalveolar lavage fluid | 14 | 15 |
|  |  | Fibrobronchoscope brush biopsy | 6 | 13 |
|  |  | Sputum | 75 | 104 |
|  |  | Nasal swabs | 5 | 8 |
|  |  | Pharyngeal swabs | 126 | 398 |
|  |  | Feces | 44 | 153 |
|  |  | Blood | 3 | 307 |
|  |  | Urine | 0 | 72 |
| Xu et al.,<br>2020 (18) | Prospective study | Nasopharyngeal swab | 22 | 49 |
|  |  | Rectal swab | 43 | 49 |
| Lo et al.,<br>2020 (16) | Prospective study | Nasopharyngeal swab | 57 | 84 |
|  |  | Sputum | 1 | 1 |
|  |  | Urine | 0 | 49 |
|  |  | Feces | 46 | 79 |
| Chan et al.,<br>2020 (19) | Case series | Nasopharyngeal swab | 4 | 5 |
|  |  | Throat swab | 2 | 3 |
|  |  | Sputum | 2 | 2 |
|  |  | Serum | 1 | 3 |
|  |  | Plasma | 0 | 4 |
|  |  | Urine | 0 | 5 |
|  |  | Feces | 0 | 4 |
| Chen et al.,<br>2020 (7) | Retrospective study | Pharyngeal swab | 65 | 167 |
|  |  | Sputum | 155 | 206 |
|  |  | Feces | 17 | 64 |
| Liu et al.,<br>2020 (5) | Cross sectional | Sputum | 28 | 57 |
|  |  | Bronchoalveolar lavage fluid | 4 | 5 |
|  |  | Nasopharyngeal swab | 1843 | 4818 |
| Wang et al.,<br>2020 (17) | Retrospective study | Nasopharyngeal swab | 134 | 706 |
|  |  | Oropharyngeal swab | 54 | 706 |

### Data synthesis for included studies

DerSimonian–Laird (DL) random effects analysis was performed to establish a summary estimate (positivity rate/proportion) by a random effects models using Open Meta-analyst software (13) and expressed using by pooled effect estimates and their 95% confidence intervals. The narrative was be written by the lead reviewer (GMB) and then checked independently by at least one other reviewer (BJN, MVM, AM). Heterogeneity in the analyzed studies was determined using I^2^ – statistic. Sub-grouping analysis was performed based on type of clinical specimen (Fig. 2) and sampling site (Fig. 3)

**Figure 2:**
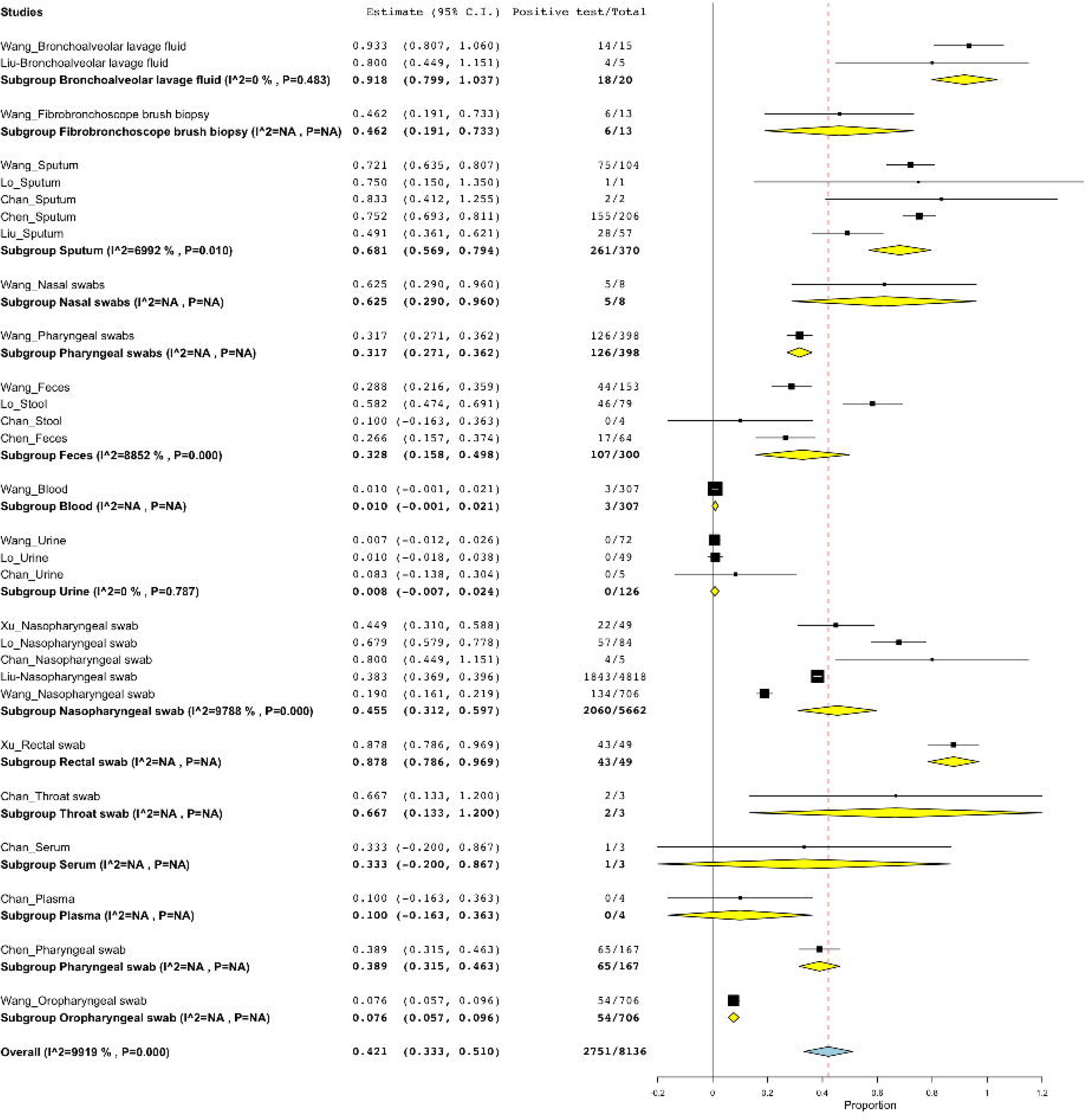
The rate of detection based on type of clinical specimen. BLF specimen had the positivity rate of 91.8% (18/20; 95%:79.9-103.7%), 87.8 % (43/49; 95%CI:78.6%-96.9%) for rectal swab, 68.1% (261/370; 95%CI:56.9%-79.4%) for sputum specimen, 62.5% (5/8; 95%CI:29.0%-96.0%) for nasal swabs, 46.2% (6/13; 95%CI:19.1%-73.3%) for FBB specimen, 31.7% (126/398; 95%CI: 27.1%-34.1%) for pharyngeal swabs, 32.8% (107/300; 95%CI:15.8%-49.8%) for feces, 1.0% (3/3017; 95%CI: -0.1%-2.1%) for blood sample, 0.8% (0/126; 95%CI:-0.7%-2.4%) for urine sample, 45.5% (2060/5662; 95%CI:31.2%-59.7%) for nasopharyngeal swab, 87.8% (43/49; 95%CI:78.6%-96.9%), 66.7% (2/3; 95%CI: 11.3%-120.0%) for throat swab, 33.3% (1/3; 95%CI:-20.0%-82.8%) for serum, 10.0% (0/4; 95%CI-16.3%-36.3%) for plasma sample, 38.9% (65/167; 95%CI:31.5%-46.3%) and 7.6% (54/706; 95%CI:5.7%-9.6%) for oropharyngeal swab.

**Figure 3:**
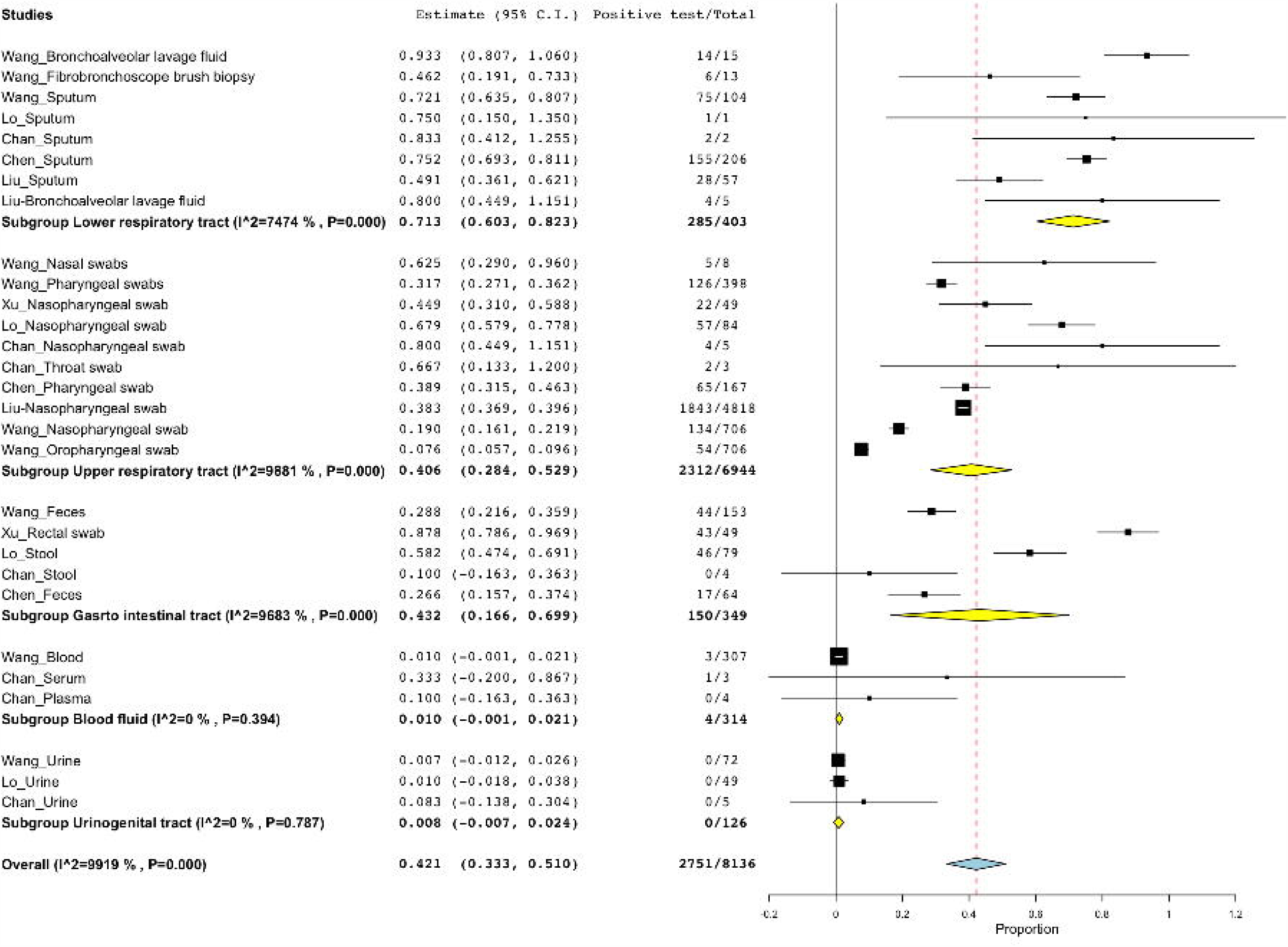
Positive detection rate based on sampling site. About 403 specimens were collected for detection of SARS-CoV-2 from LRT where 71.3% (285/403; 95%CI:60.3%-82.3%) tested positive. Specimens from URT had the positivity rate of 40.6% (2312/6944; 95%CI:28.4%-52.9%). Gastro intestinal tract (GIT) specimens recorded 43.2% positive SARS-CoV-2 (150/349; 95%CI:16.6%-69.9%). Blood fluids and UGT had the positivity rate of 1.0% (4/314; 95%CI:-0.1%-2.1%) and 0.8% (0/126; 95%CI:-0.7-2.4%), respectively.

### Quality assessment

Output generated Open Meta Analyst software through cumulative forest plot was indirectly employed to assess the publication bias (14). As such, the estimate of the proportions decreased with the increase in sample size. This decrease, could be due to publication bias or it could be due to small-study effects. However, when analysis was limited to study with at least 100 sample size, the overall positive detection rate would have been 34.6% (95%CI, 17.9%-51.3%) while that with all studies included regardless of the sample size was 42.7% (95%CI: 32.2-53.3%) (Additional file 2). On the other hand, the quality of the included studies (risk of bias) was assessed using Newcastle Ottawa Scale adapted for cross sectional studies (Additional file 3) as previously described elsewhere (15). The risk of bias was evaluated by two independent reviewers (GMB & BJN). Discrepancies were resolved by consensus, and/or consulting a third reviewer (MVM) where necessary.

## Results

### Characteristics of the included studies

Of 780 pooled studies, 7 studies qualified for final analysis (Table 1). Among the seven articles, information reported on positive detection rate were extracted from four studies (5,6,16,17) for suspected cases, while in 3 studies (7,18,19) information were recorded if at least one specimen tested positive (confirmed cases) from the simultaneous tested specimens and the recording of positive tests were stopped when at least 2 consecutives qTR-PCR tested negative in all specimens (7,18). Different types of clinical specimens were tested for SARS-CoV-2, bronchoalveolar lavage fluid (BLF) (5,6), fibrobronchoscope brush biopsy (FBB) (6), sputum (5–7,16,17,19). Some of the studies reported pharyngeal specimen without specifying whether the route was nasal of oral (6,7). One study reported nasal and pharyngeal swabs combined, but for the purpose of this review, it was categorized as nasopharyngeal swabs (5). Nasal swabs were reported in one study (6), feces (6,7), blood (6), urine (6,16,19), nasopharyngeal (5,16–19), sputum (5,7,19), rectal swab (18), oropharyngeal (17), serum and plasma (19).

### Positive detection rates for different types of clinical specimens

A total of 8136 specimens of the suspected cases were tested for SARS-CoV-2, where majority of the specimens were nasopharyngeal swabs (69.6%; 5662/8136), with only three specimens for throat swabs and serum sample. Regarding the sampling site, most of the specimens were collected from the upper respiratory tract (URT) (85.3%; 6944/8136) whereas few specimens were reported from urinogenital tract (UGT) (1.5%; 126/8126). There was high positive rate of 91.8% (18/20; 95%:79.9-103.7%) and 71.3% (285/403; 95%CI:60.3%-82.3%) for BLF clinical specimens and LRT site, respectively. While low detection rate of 0.8% (0/126; 95%CI:-0.7%-2.4%) for urine sample and 0.8% (0/126; 95%CI:-0.7-2.4%) for specimens collected from UGT.

## Discussion

For the purpose of guiding the selection of clinical specimens for clinical diagnosis of COVID-19, conducting the current systematic review which aimed at profiling the positive detection rate from different clinical specimens using PCR was necessary. In this study, SARS-CoV-2 was detected in specimens from 8136 specimens of patients suspected and/ or confirmed with COVID-19, with LRT specimens being the most effective in detecting the virus by 71.3%. There was no evidence of detecting virus from the urinogenital specimens.

Regarding the type of clinical specimens, BLF had the positivity rate of 91.8% followed by rectal swab (87.8 %) then sputum specimens (62.5%). Nasopharyngeal swab which is commonly and widely used (1) had a positive detection rate of 45.5%. Low detection rate was observed in oropharyngeal swab (7.6%) with zero detection from urine samples. There was low detection of SARS-CoV-2 in blood (1.0%) blood sample, but moderate in serum (33.3%) and plasma (10.0%). More importantly, the virus was detected in 32.8% of feces, suggesting that SARS-CoV-2 resist the acidic medium of the human gut and can be transmitted by the fecal route. In support fecal route transmission, Wang et al (6) reported live virus form the stool specimens.

Majority of the specimens were nasopharyngeal swabs (69.6%). This is in-line with the study which reported the clinical characteristics of patients with COVID-19 where nasopharyngeal swab was typically used to confirm the diagnosis (20). Sampling of nasopharyngeal swab is less invasive when compared to other specimens such as BLF (21) which makes it more preferable sample. Sputum another LRT samples was found to have a good detection rate but dry cough as common clinical presentation for COVID-19 patients limit its availability (22). On the other hand, rectal swab was the second in recording high positive rate exceeding even the URT specimens such as sputum. With regards, to rectal swab can be considered as a representative of gastro intestinal specimen.

This study was limited from the symptoms suggestive for COVID-19 in four studies (5,6,16,17), hence limiting the determination of true negative which had an impact on the denominator (total samples tested). However, in 3 studies (7,18,19) information were recorded if at least one specimen tested positive (confirmed cases) from the simultaneous tested specimens and the recording of positive tests were stopped when at least 2 consecutives qTR-PCR tested negative in all specimens (7,18). In addition, this review was limited to poor quality of the study design and small sample size for some of the investigated specimens, i.e., BLF and rectal swabs.

## Conclusion

In this study, SARS-CoV-2 was highly detected in lower respiratory tract, with zero detection of virus from the urogenital specimens. Regarding the type of clinical specimens, bronchoalveolar lavage fluid had the highest positive rate followed by rectal swab then sputum specimens. Nasopharyngeal swab which is widely used had a moderate detection rate. Low positive rate was recorded in oropharyngeal swab and blood sample while no virus was found in urine samples. More importantly, the virus was detected in feces, suggesting SARS-CoV-2 transmission by the fecal route. The use of specimens such as bronchoalveolar lavage fluid, sputum and rectal swab is recommended for clinical diagnosis of COVID-19.

## Data Availability

All relevant data are within the manuscript and its Supporting Information files (Additional file 1, 2 &3).

## Acknowledgment

Authors, thank Deodatus Sabas, Librarian at Muhimbili University of Health and Allied Sciences, Dar es Salaam (MUHAS), Tanzania for his support during literature search. Authors acknowledge the training support received from MUHAS through Systematic Review and Meta-analysis workshop funded by Swedish International Development Cooperation Agency, Sweden.

## Authors contributions

Conceptualization, Data curation, Formal analysis, Methodology, Manuscript drafting: George M. Bwire.

Data curation, Formal analysis, Methodology, Manuscript review and editing: Belinda J. Njiro.

Formal analysis and Manuscript review and editing: Mtebe V. Majigo

Methodology and Manuscript review and editing: Akili Mawazo

All authors have read and approved the final version of manuscript.

## Competing interest statement

Authors declare that, they have competing interest

## Ethics statement

Not applicable

## Funding

The authors received no specific funding for this work.

## TABLES AND FIGURES LEGENDS

**Additional file 1:** PubMed search results

**Additional file 2:** Cumulative forest plot for assessing the publication bias

**Additional file 3:** Assessment of risk of bias using Newcastle-Ottawa Scale adapted for cross-sectional studies

## Notes

### Competing Interest Statement

The authors have declared no competing interest.

### Author Declarations

The type of study (Systematic review) does not require ethical clearance.

